# The impact of COVID-19 on the reproductive health of people living in Australia: findings from an online survey

**DOI:** 10.1101/2020.08.10.20172163

**Authors:** Jacqueline Coombe, Fabian Kong, Helen Bittleston, Hennie Williams, Jane Tomnay, Alaina Vaisey, Sue Malta, Jane Goller, Meredith Temple-Smith, Louise Bourchier, Andrew Lau, Jane S Hocking

## Abstract

**Introduction:** Australia introduced ‘lockdown’ measures to control COVID-19 on 22 March 2020. For two months, Australians were asked to remain at home and only leave for essential activities. We investigate the impact this had on sexual and reproductive health (SRH).

**Methods:** Australians aged 18+ were eligible to participate in an online survey from 23 April-11 May 2020. Questions included contraceptive use, pregnancy intentions and access to SRH services. We report on the experiences of 518 female participants aged <50 years. Pregnancy intentions and contraceptive use were analysed using descriptive statistics. Odds ratios and 95% confidence intervals were calculated to investigate difficulty accessing SRH products and services. Qualitative data were analysed using descriptive thematic analysis.

**Results:** Most participants (55.4%, 287/518) were aged 18-24 years. Most (76.1%, 379/498) indicated they were trying to avoid pregnancy. The oral contraceptive pill was the most common single method used (20.8%; 107/514). Nearly 20% (101/514) reported they were not using contraception. Older women (OR=0.4; 95%CI: 0.1, 0.9 for 25-34 vs 18-24 years) and those employed (OR=0.4; 95%CI: 0.2, 0.7) had less trouble accessing contraception during lockdown. Women aged 25-34 (OR=0.4; 95%CI: 0.3, 0.7) or 35-49 years (OR=0.3; 95%CI: 0.1, 0.6) were less likely to experience difficulty accessing feminine hygiene products. Qualitative analysis suggested that COVID-19 affected pregnancy plans, with participants delaying childbearing, or deciding to remain childfree.

**Conclusion:** COVID-19 lockdown impacted the SRH of Australian women. Findings highlight the importance of continued access to SRH services and products during global emergencies.

**KEY MESSAGES:**

- Nearly a third of participants reported difficulties accessing their usual feminine hygiene products during lockdown in Australia.
- Participants reported delaying childbearing or deciding to remain childfree due to the COVID-19 pandemic.
- Ensuring continued access to sexual and reproductive health services and products for all who require them during global emergencies is essential.

## INTRODUCTION

In response to rising COVID-19 cases, the Australian Government implemented a country-wide lockdown extending from 22 March to 8 May 2020. During this time, people were asked to remain in their homes and only leave to access essential goods and services, to receive or provide care, to exercise, or to attend work or education where these activities could not take place at home. This approach was largely successful; at the end of the lockdown period, Australia had recorded 6,914 cases and 97 deaths[1], numbers that pale in comparison with those recorded elsewhere[2]. However, Australia is not immune to the social and economic hardships the pandemic caused, nor the impact on the provision of and access to sexual and reproductive health (SRH).

In March, one of the largest condom producers in the world warned of a global shortage[3], potentially impacting the sexual health of millions. Domestic travel restrictions in Australia exacerbated an already complicated abortion care system with patients and providers unable to travel to seek or provide care[4]. Marie Stopes Australia warned of an increased risk of unplanned pregnancy and sexually transmissible infections (STIs) as contraception and abortion, as well as pregnancy and STI tests become less accessible[4]. Plan International reported on the impact on menstrual hygiene, highlighting the significant impact of the pandemic on the ability of menstruating people to manage their periods safely and hygienically[5]. News outlets reported predictions of a falling fertility rate as couples reassessed desires to reproduce during a health crisis and amidst the resulting social and economic hardships[6].

We are conducting serial cross-sectional surveys to investigate the impact of COVID-19 on the SRH of people living in Australia. In this paper, we report on the results from the first survey and explore the impact of Australia-wide lockdown on reproductive health including pregnancy intentions and contraception access.

## METHODS

Data reported here were collected as part of the Sexual and Reproductive Health during COVID-19 survey open from 23 April-11 May 2020. People aged 18+ who were living in Australia were eligible to participate. Repeat waves will be conducted throughout 2020, and a cohort analysis undertaken for participants who respond to multiple surveys. The survey comprises three sections: demographics, sexual practices and reproductive health. In this paper we report on data pertaining only to the baseline survey and for those participants who completed the reproductive health section. The study was approved by the University of Melbourne Human Research Ethics Committee (ID: 2056693).

### Recruitment

Recruitment flyers were distributed to SRH newsletter lists, emailed to colleagues, posted on student noticeboards and social media. Paid Facebook ads were also utilised. Interested participants clicked on a link which took them to a plain language statement. Participants were asked to consent to participate prior to commencing the survey.

### Data collection

Participants were asked about changes in sexual practices, intimate relationships, contraceptive use and access, pregnancy intention and access to and use of SRH services. Several questions investigated whether during lockdown, participants had difficulty accessing contraception, SRH products like pregnancy tests, lubrication and feminine hygiene products. They were also asked about their healthcare access during lockdown, including whether restrictions had impacted their ability to access emergency contraception or termination of pregnancy. Free-text responses to a question regarding the impact of COVID-19 on pregnancy plans were also collected.

### Data analysis

Analysis was limited to reproductive aged women (<50 years). Descriptive statistics were used to describe socio-demographic characteristics, pregnancy intentions and contraception use. Odds ratios and 95% confidence intervals were calculated to investigate factors associated with healthcare access and difficulty in accessing contraception, SRH products and feminine hygiene products. As this was an explorative analysis, regression modelling to examine the impact of confounding on associations was not undertaken. Chi square tests and t tests (where appropriate) were used to investigate factors associated with providing free-text responses. As not everyone completed all questions, missing data were excluded, but denominators are provided to place missing data in context. Responses to the free-text question were imported into NVivo qualitative analysis software [7]. Utilising thematic analysis [8], we took a largely descriptive and semantic approach to the data, looking for the overt meaning of the comments.

### Patient and Public Involvement Statement

Patients and the public were not involved in the development of this study; however, the survey was pilot tested with several people not involved with the study and their feedback incorporated into the final survey. A lay summary will be available to participants on our website[9] upon publication of our results.

## RESULTS

### Demographics

A total of 1187 people consented to participate of whom 965 (81.3%) answered the questions on sexual practices and 625 (52.6%) answered questions on reproductive health. Of these 625, 518 (82.9%) reported that they were female and aged <50 years and were included in the analysis. There was no difference in average age between those who completed the reproductive health questions and those who did not (25.7 vs 27.0 years, p=0.12). Among participating women, 55.4% (287/518) were aged 18-24 years, 36.7% (397/518) were in a cohabitating relationship and 32.2% (167/518) reported being single. Overall, 397 (77.2%) were living in urban areas of Australia and 61.1% (315/516) were employed at the time of the survey. A small proportion reported that they or their partner was pregnant (1.9%, 10/516) and most (76.1%, 379/498) indicated that they were trying to avoid pregnancy. Nearly 20% (101/514) reported they were not using any contraception, with the oral contraceptive pill being the most common single method used (20.8%; 107/514) and condoms and the pill (6.4%; 33/514) being the most common dual method used (Table 1).

**Table 1.**
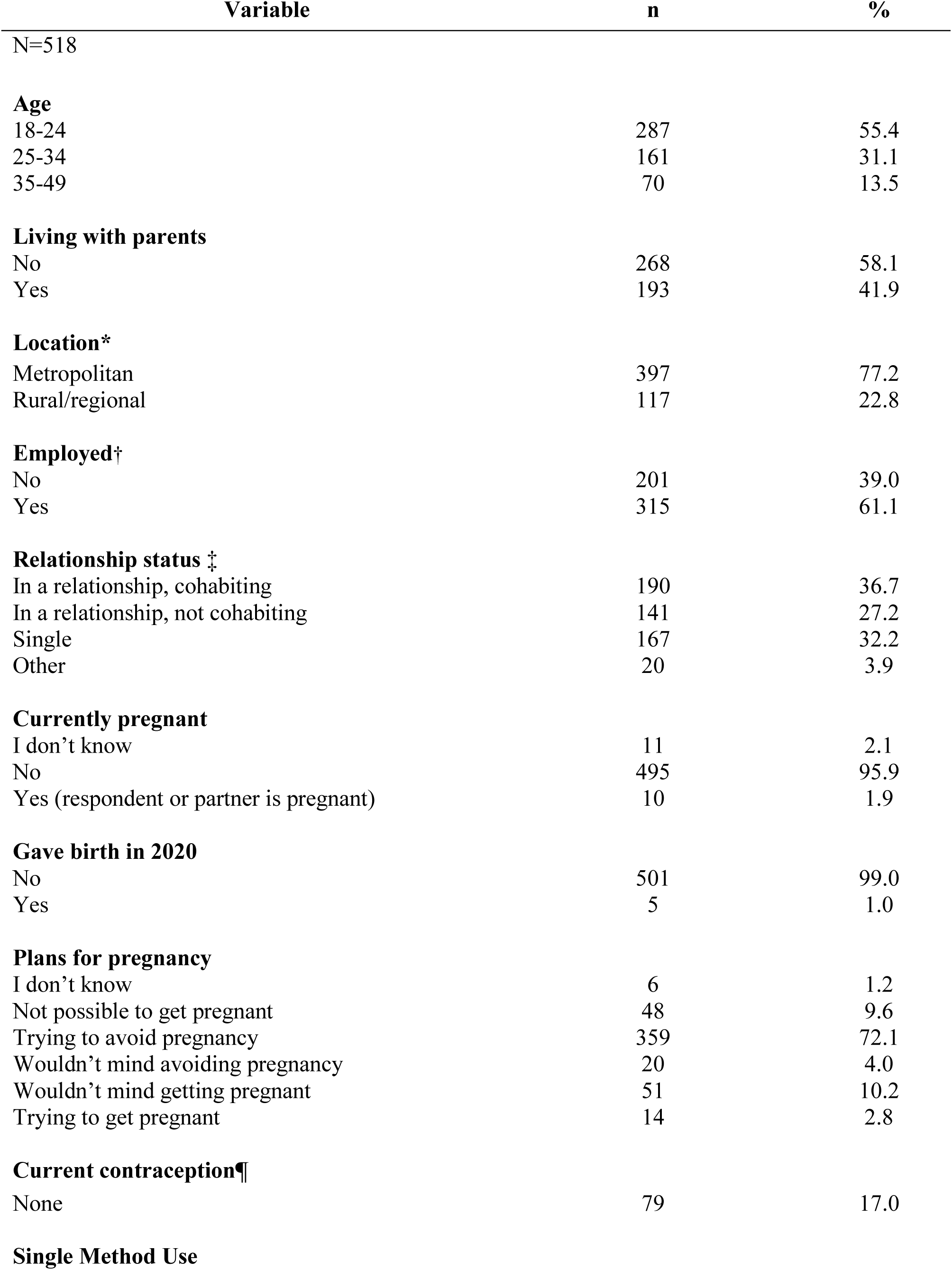
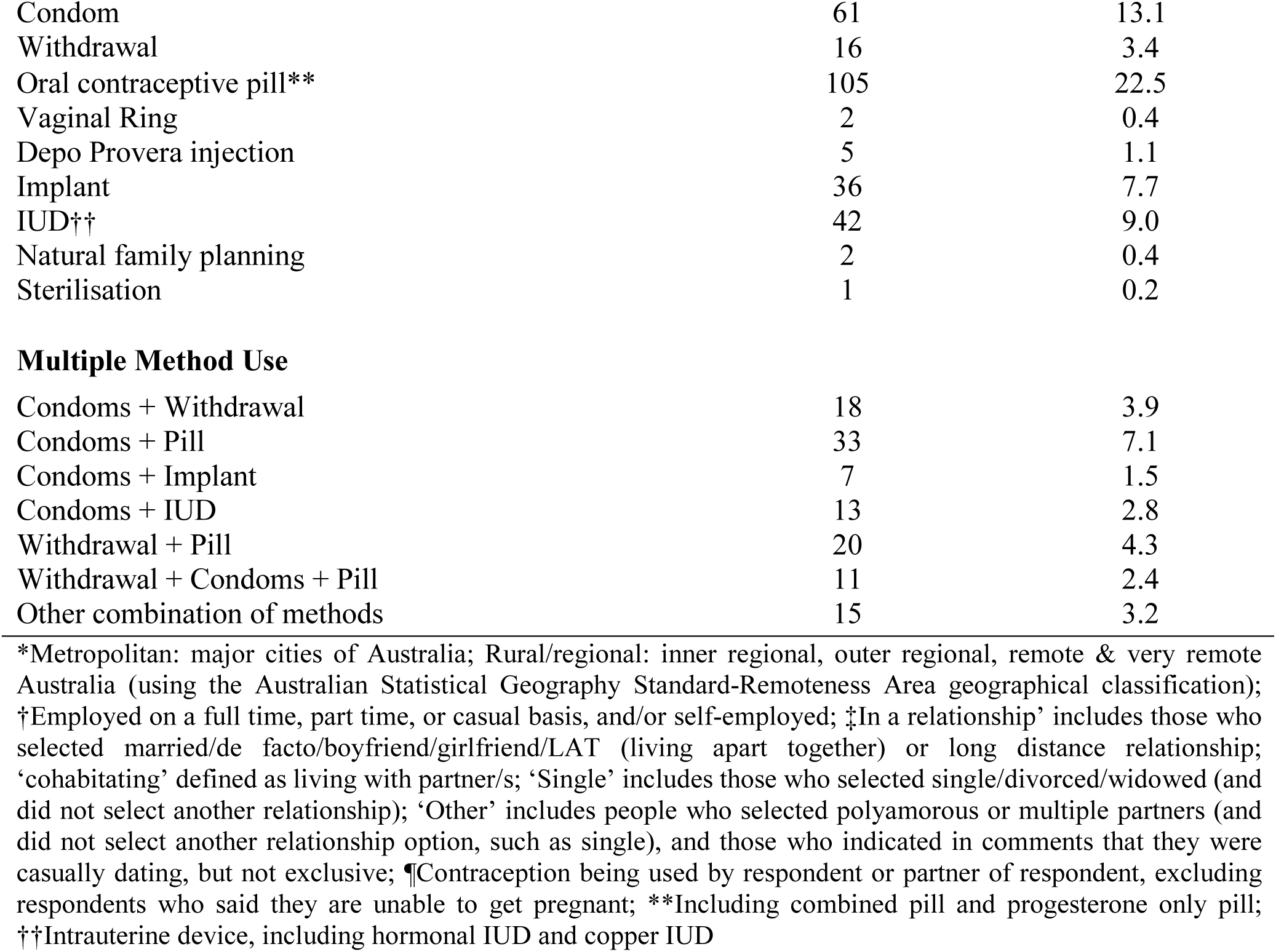
Sociodemographic characteristics, contraceptive use and pregnancy intentions of 518 female survey participants

| Variable | n | % |
| --- | --- | --- |
| N=518 |  |  |
| **Age** |  |  |
| 18-24 | 287 | 55.4 |
| 25-34 | 161 | 31.1 |
| 35-49 | 70 | 13.5 |
| **Living with parents** |  |  |
| No | 268 | 58.1 |
| Yes | 193 | 41.9 |
| **Location\*** |  |  |
| Metropolitan | 397 | 77.2 |
| Rural/regional | 117 | 22.8 |
| **Employed†** |  |  |
| No | 201 | 39.0 |
| Yes | 315 | 61.1 |
| **Relationship status ‡** |  |  |
| In a relationship, cohabiting | 190 | 36.7 |
| In a relationship, not cohabiting | 141 | 27.2 |
| Single | 167 | 32.2 |
| Other | 20 | 3.9 |
| **Currently pregnant** |  |  |
| I don't know | 11 | 2.1 |
| No | 495 | 95.9 |
| Yes (respondent or partner is pregnant) | 10 | 1.9 |
| **Gave birth in 2020** |  |  |
| No | 501 | 99.0 |
| Yes | 5 | 1.0 |
| **Plans for pregnancy** |  |  |
| I don't know | 6 | 1.2 |
| Not possible to get pregnant | 48 | 9.6 |
| Trying to avoid pregnancy | 359 | 72.1 |
| Wouldn't mind avoiding pregnancy | 20 | 4.0 |
| Wouldn't mind getting pregnant | 51 | 10.2 |
| Trying to get pregnant | 14 | 2.8 |
| **Current contraception¶** |  |  |
| None | 79 | 17.0 |
| **Single Method Use** |  |  |
| Condom | 61 | 13.1 |
| Withdrawal | 16 | 3.4 |
| Oral contraceptive pill** | 105 | 22.5 |
| Vaginal Ring | 2 | 0.4 |
| Depo Provera injection | 5 | 1.1 |
| Implant | 36 | 7.7 |
| IUD†† | 42 | 9.0 |
| Natural family planning | 2 | 0.4 |
| Sterilisation | 1 | 0.2 |
| Multiple Method Use |  |  |
| Condoms + Withdrawal | 18 | 3.9 |
| Condoms + Pill | 33 | 7.1 |
| Condoms + Implant | 7 | 1.5 |
| Condoms + IUD | 13 | 2.8 |
| Withdrawal + Pill | 20 | 4.3 |
| Withdrawal + Condoms + Pill | 11 | 2.4 |
| Other combination of methods | 15 | 3.2 |

\*Metropolitan: major cities of Australia; Rural/regional: inner regional, outer regional, remote & very remote Australia (using the Australian Statistical Geography Standard-Remoteness Area geographical classification); †Employed on a full time, part time, or casual basis, and/or self-employed; ‡In a relationship' includes those who selected married/de facto/boyfriend/girlfriend/LAT (living apart together) or long distance relationship; 'cohabitating' defined as living with partner/s; 'Single' includes those who selected single/divorced/widowed (and did not select another relationship); 'Other' includes people who selected polyamorous or multiple partners (and did not select another relationship option, such as single), and those who indicated in comments that they were casually dating, but not exclusive; ¶Contraception being used by respondent or partner of respondent, excluding respondents who said they are unable to get pregnant; \*\*Including combined pill and progesterone only pill; ††Intrauterine device, including hormonal IUD and copper IUD

When asked about the ease of access to SRH products and services during lockdown, 9.2% (37/404) reported that they had trouble accessing contraception, 4.6% (13/280) had trouble accessing SRH products like pregnancy tests and lubrication and 32.7% (134/410) had trouble accessing their usual feminine hygiene products. Of those who experienced difficulty accessing feminine hygiene products, nearly half (48.5%, 65/134) said that they changed their use of products as a consequence.

Univariate analysis found that women aged 25-34 years (OR=0.4; 95%CI 0.1, 0.9) compared with women aged 18-24 years and those employed (OR=0.4; 95%CI 0.2, 0.7) had less trouble accessing contraception during lockdown. No factors were associated with accessing SRH products. Women aged 25-34 years (OR=0.4; 95%CI 0.3, 0.7) or 35-49 years (OR=0.3; 95%CI 0.1, 0.6) were less likely to experience difficulty accessing feminine hygiene products than those aged 18-24 years. Those in a non-cohabitating relationship (OR=1.9; 95%CI 1.1, 3.1) and those living with parents (OR=2.4; 95%CI 1.5, 3.8) were more likely to have difficulty accessing feminine hygiene products than those in a cohabitating relationship (Table 2).

**Table 2.**
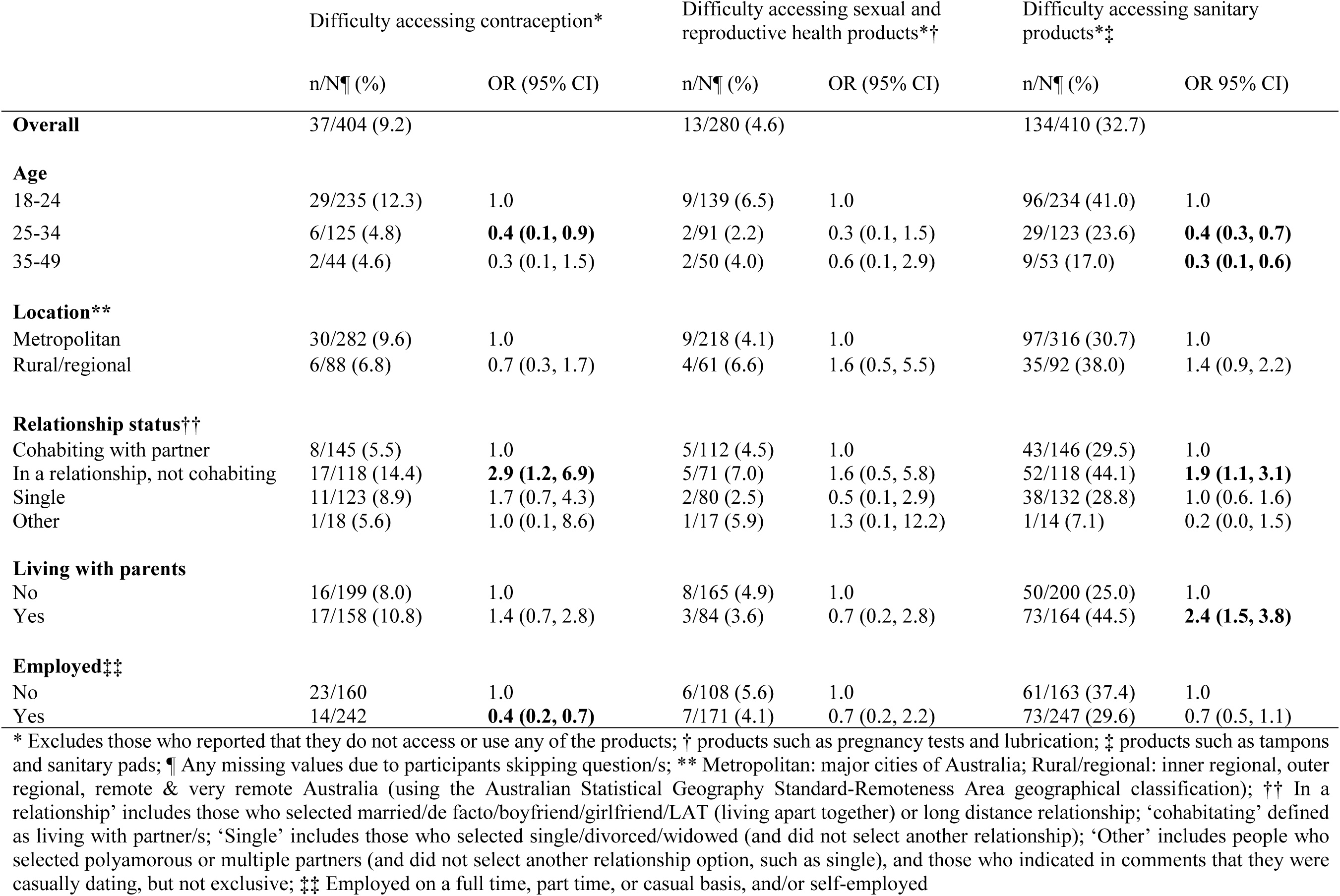
Access to sexual and reproductive health products

When asked about their access to health services, 22.4% (114/510) reported that they had needed to access healthcare services for reasons related to their sexual or reproductive health during lockdown, 33.1% (165/498) had accessed an online health service (although this may have been for reasons other than SRH), and 2.4% (12/510) had needed to access emergency contraception or termination of pregnancy services for themselves, or for their partner. There were no significant differences between women living in metro versus rural/regional areas.

Univariate analysis found that women aged 25-34 years (OR=1.6; 95%CI: 1.0, 2.5 compared with women aged 18-24 years were more likely to have accessed healthcare for their SRH reasons, and single women (OR=0.6; 95%CI: 0.3, 1.0) were less likely to have accessed services compared to women in a cohabitating relationship. Women aged 25-34 years were more likely to have accessed online healthcare services than women aged 18-24 (OR=1.6, 95%CI: 1.0, 2.4). No factors were associated with accessing emergency contraception or termination of pregnancy services (Table 3).

**Table 3.** Access to health services

|  | Accessed termination of pregnancy service* |  | Accessed sexual and reproductive healthcare services† |  | Accessed online healthcare/telehealth services‡ |  |
| --- | --- | --- | --- | --- | --- | --- |
|  | n/N¶ (%) | OR (95% CI) | n/N¶ (%) | OR (95% CI) | n/N¶ (%) | OR 95% CI |
| **Overall** | 12/510 (2.4) |  | 114/510 (22.4) |  | 165/498 (33.1) |  |
| **Age** |  |  |  |  |  |  |
| 18-24 | 10/283 (3.5) | 1.0 | 57/282 (20.2) | 1.0 | 80/274 (29.2) | 1.0 |
| 25-34 | 2/157 (1.3) | 0.4 (0.1, 1.6) | 45/158 (28.5) | **1.6 (1.0, 2.5)** | 61/156 (39.1) | **1.6 (1.0, 2.4)** |
| 35-49 | 0/70 (0.0) | - | 12/70 (17.1) | 0.8 (0.4, 1.6) | 24/68 (35.3) | 1.3 (0.8, 2.3) |
| **Location\*\*** |  |  |  |  |  |  |
| Metropolitan | 8/394 (2.0) | 1.0 | 89/394 (22.6) | 1.0 | 124/386 (32.1) | 1.0 |
| Rural/regional | 4/112 (3.6) | 1.8 (0.5, 6.0) | 25/112 (22.3) | 1.0 (0.6, 1.6) | 39/108 (36.1) | 1.2 (0.8, 1.9) |
| **Relationship status††** |  |  |  |  |  |  |
| Cohabiting with partner | 2/187 (1.1) | 1.0 | 47/188 (25.0) | 1.0 | 60/184 (32.6) | 1.0 |
| In a relationship, not cohabiting | 5/138 (3.6) | 3.5 (0.7, 18.2) | 31/137 (22.6) | 0.9 (0.5, 1.5) | 39/133 (29.3) | 0.9 (0.5, 1.4) |
| Single | 5/165 (3.0) | 2.9 (0.6, 15.1) | 37/165 (16.4) | **0.6 (0.3, 1.0)** | 57/161 (35.4) | 1.1 (0.7, 1.8) |
| Other | 0/20 (0.0) | - | 9/20 (45.0) | 2.5 (1.0, 6.3) | 9/20 (45.0) | 1.7 (0.7, 4.3) |
| **Living with parents** |  |  |  |  |  |  |
| No | 3/264 (1.1) | 1.0 | 60/265 (22.6) | 1.0 | 89/259 (34.4) | 1.0 |
| Yes | 6/189 (3.2) | 2.9 (0.7, 11.6) | 38/188 (20.2) | 0.9 (0.5, 1.4) | 53/184 (28.8) | 0.8 (0.5, 1.2) |
| **Employed‡‡** |  |  |  |  |  |  |
| No | 4/197 (2.0) | 1.0 | 41/196 (20.9) | 1.0 | 63/190 (33.2) | 1.0 |
| Yes | 8/311 (2.6) | 1.3 (0.4, 4.3) | 73/312 (23.4) | 1.2 (0.7, 1.8) | 101/306 (33.0) | 1.0 (0.7, 1.5) |

\*including abortion services and emergency contraception; †Including general practice, specialist sexual or reproductive health provider, pharmacy, hospital, and counselling/psychology services; ‡Including online pharmacy, online STI testing, and seeing a general practitioner via telephone/internet (telehealth); ¶Any missing values due to participants skipping question/s; \*\* Metropolitan: major cities of Australia; Rural/regional: inner regional, outer regional, remote & very remote Australia (using the Australian Statistical Geography Standard-Remoteness Area geographical classification); †† In a relationship' includes those who selected married/de facto/boyfriend/girlfriend/LAT (living apart together) or long distance relationship; 'cohabitating' defined as living with partner/s; 'Single' includes those who selected single/divorced/widowed (and did not select another relationship); 'Other' includes people who selected polyamorous or multiple
partners (and did not select another relationship option, such as single), and those who indicated in comments that they were casually dating, but not exclusive; ‡‡ Employed on a full time, part time, or casual basis, and/or self-employed

### The impact of COVID-19 on plans for pregnancy

We also explored the impact of COVID-19 on plans for pregnancy, and participants were asked to respond to a free-text question about this *(Has the COVID-19 (coronavirus) pandemic impacted your future plans for pregnancy? Please tell us about it in the box below)*. In total, 217 participants provided a comment. Participants who chose to respond to the free-text question were older than those who did not provide a response (26.8 years vs. 25.1 years; p<0.01). They were also more likely to be in a cohabitating relationship than those who did not provide a response (47.9% vs. 28.4%; p<0.01). Participants who responded to the free-text question were more likely to be trying to get pregnant (6.5% vs 0.0%; p<0.01), or at a stage where they did not mind getting pregnant (17.6% vs 4.6%; p<0.01) than those who did not respond to the free text question. Participant responses largely fell into one of four key themes. These themes are presented below, and illustrative quotes can be found in Table 4.

**Table 4.**
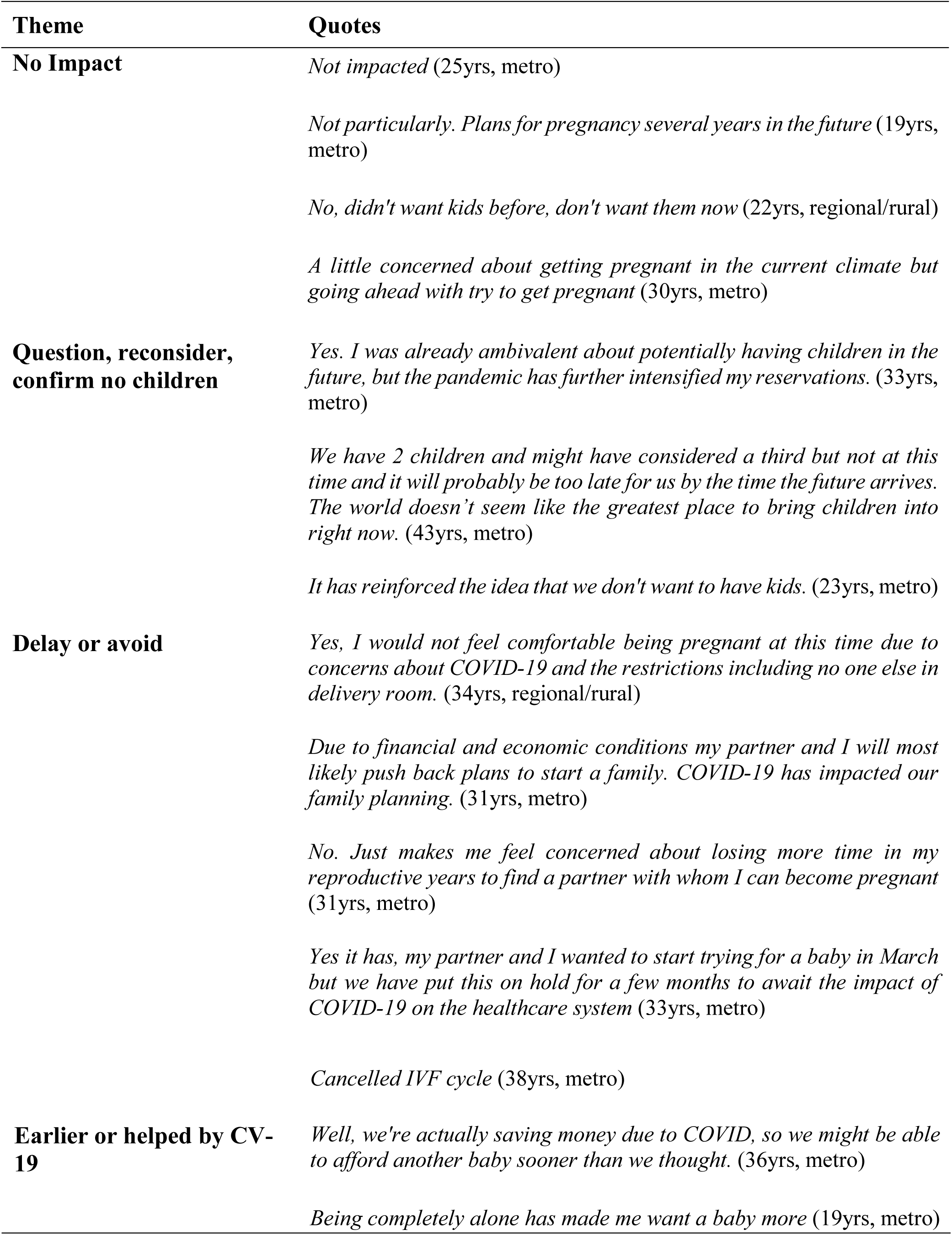
Illustrative quotes for themes about the impact of COVID-19 on pregnancy plans

#### Theme 1: No impact

Most participants reported no impact on their future plans for pregnancy, reporting variations of ‘no’ or ‘no impact’. Others were more expansive in their responses, noting that COVID-19 has not changed their plans for pregnancy, largely because these plans are in the distant, hopefully COVID free future, or because they do not want to have children. For some, COVID-19 had not impacted plans to conceive, with a few indicating that they will continue to try to conceive this year.

#### Theme 2: COVID-19 has led to questioning, or reconsidering decisions to have children, or confirmed desire not to have children

For others, the pandemic had led them to question whether to have children. Many of these participants indicated pre-COVID reservations about having children, with the pandemic leading to further reservations. For others, the pandemic solidified their decision not to have children at all. These comments were different to those reported in ‘no impact’, in that they clearly linked their desire not to have children to the pandemic and the general ‘state of the world’.

#### Theme 3: Delay or avoid pregnancy due to COVID-19

Some participants reported delaying or avoiding pregnancy due to COVID-19. These participants cited concerns about pregnancy care during the pandemic, not putting undue strain on the healthcare system, and concerns about the impact of the virus on pregnant women and newborns. Other participants reported a delay in plans for pregnancy due to their changing life circumstances. Although these participants had not planned to conceive in 2020, because of delayed travel, delayed marriage, career interruptions, changing financial situations, and an inability to meet a partner due to lockdown restrictions, these plans are delayed further into the future and, for some, indefinitely. Finally, a group of participants reported that earlier in the year they had been actively trying to conceive or were planning to conceive this year. These participants reported putting these plans on hold, either because they were unable to continue trying to conceive due to the cancellation of IVF and other reproductive services, or because they had made the decision to stop actively trying during the pandemic.

#### Theme 4: Pregnancy plans earlier, or helped by COVID-19

For a small number of participants, COVID-19 had provided favourable circumstances for conception. For some changing life plans, like an inability to travel overseas, had led them to move their plans to conceive earlier. Others reported being able to save more money due to the COVID-19 restrictions, also facilitating earlier conception than otherwise planned. In contrast to those questioning or deciding not to have children due to the pandemic, some participants said that the pandemic had increased their desire for children. These comments were often framed within the isolation of lockdown.

## DISCUSSION

The COVID-19 pandemic and the resulting restrictions have clearly impacted the ability of Australian women to access reproductive healthcare and products. While most participants reported little difficulty in accessing their usual contraceptive methods and appeared to continue to access healthcare as needed, nearly a third reported difficulties accessing their usual feminine hygiene products. Many also reported delaying plans for pregnancy due to the pandemic. Although our findings should be considered within their limitations, namely the homogeneity of our sample, our findings offer unique insight into the SRH impact of COVID-19 on Australian women.

Our findings show that most participants were able to continue to access their usual contraceptive method during the first lockdown in Australia. However, of those reporting difficulties accessing their usual contraceptive method, younger women, and women who were unemployed were more likely to report difficulties. Recent analysis examining those hardest hit by the economic fall-out from COVID-19 show that women and young people are most adversely affected, largely due to their disproportionate representation in industries adversely impacted by the pandemic to-date[10]. While there are many contraceptive options available on the Australian market, the most effective methods require a prescription by a healthcare provider and out-of-pocket costs from the user to purchase[11]. While switching to a different, potentially cheaper method sounds feasible, in practice, women often spend a significant amount of time finding a suitable contraceptive method, and even switching between different brands of the oral contraceptive pill can bring new, unwanted side effects[12].

Accessing one’s preferred contraceptive method is also essential to the prevention of unintended pregnancy. Most participants in our study reported intentions to avoid pregnancy, with few reporting actively trying to conceive. Importantly, many reported delaying or indefinitely putting plans for pregnancy on hold due to the pandemic. The impact of COVID-19 on pregnant and birthing women has been profound. While the limited research thus far suggests that pregnant women are not at increased risk of contracting COVID-19 or experiencing more severe symptoms[13], the increased stress and anxiety experienced by these women as they navigate care under strict COVID-19 regulations is becoming clear[13-15]. It is perhaps unsurprising then that women are delaying plans for pregnancy in the current environment. What impact this might have on the Australian fertility rate in the future is unclear although, at least for the participants in our study, it seems that a post lockdown baby-boom is unlikely.

Finally, nearly a third of participants who reported usually accessing feminine hygiene products reported difficulties in accessing their usual products, and nearly half of these reported changing their use as a result. During the first phase of the pandemic Australians were repeatedly reassured by the Australian Government and large supermarkets that there was no shortage of supplies of essential goods, and that the lack of toilet paper, tissues, flour, eggs and pasta (among other things) was a consequence of the panic buying that typified the first phase of the pandemic. This behaviour did not go unnoticed by the Prime Minister, who asked Australian’s to “just stop it”[16]. While this shortage may indeed have been inadvertently caused by the panic buying of panicked Australian’s, the lack of feminine hygiene products, combined with difficulties accessing contraception and delayed plans to conceive are clear examples of the gendered impact of COVID-19. Although evidence regarding the gendered impact of the pandemic, including but not limited to SRH is mounting, the Australian Government continues to largely ignore this in their pandemic response. Recent examples include changes to telehealth services that now require patients to have an existing relationship with a general practitioner providing the service[17], a move widely criticised by the SRH sector[18], and the removal of financial support for the childcare sector[19], the only sector to have their support removed thus far and a move that clearly disproportionally impacts women[20]. The COVID-19 pandemic is clearly impacting the SRH of women in Australia. Availability of all forms of contraception and SRH products and services are essential for reproductive planning, and it is vital that all who need these services continue to be able to access them as the pandemic continues.

## Data Availability

No data are available.

## Acknowledgments

We would like to thank everyone who generously gave their time to complete our survey.

## Author contribution

All authors contributed to the design and development of the survey. JC was responsible for administering the survey. HB, JH and JC conducted the analysis. JC, HB and JH interpreted the results and drafted the manuscript. All authors contributed to the revision of draft iterations of the manuscript prior to submission.

---

STROBE Statement—Checklist of items that should be included in reports of ***cross-sectional studies***

|  | Item No | Recommendation | Page No |
| --- | --- | --- | --- |
| Title and abstract | 1 | (a) Indicate the study’s design with a commonly used term in the title or the abstract | 1 |
|  |  | (b) Provide in the abstract an informative and balanced summary of what was done and what was found | 2 |
| Introduction |  |  |  |
| Background/rationale | 2 | Explain the scientific background and rationale for the investigation being reported | 3 |
| Objectives | 3 | State specific objectives, including any prespecified hypotheses | 3 |
| Methods |  |  |  |
| Study design | 4 | Present key elements of study design early in the paper | 3 |
| Setting | 5 | Describe the setting, locations, and relevant dates, including periods of recruitment, exposure, follow-up, and data collection | 3 |
| Participants | 6 | (a) Give the eligibility criteria, and the sources and methods of selection of participants | 3 |
| Variables | 7 | Clearly define all outcomes, exposures, predictors, potential confounders, and effect modifiers. Give diagnostic criteria, if applicable | 3-4 |
| Data sources/<br>measurement | 8* | For each variable of interest, give sources of data and details of methods of assessment (measurement). Describe comparability of assessment methods if there is more than one group | 3-4 |
| Bias | 9 | Describe any efforts to address potential sources of bias | 3-4 |
| Study size | 10 | Explain how the study size was arrived at | 3-4 |
| Quantitative variables | 11 | Explain how quantitative variables were handled in the analyses. If applicable, describe which groupings were chosen and why | 3-4 |
| Statistical methods | 12 | (a) Describe all statistical methods, including those used to control for confounding | 3-4 |
|  |  | (b) Describe any methods used to examine subgroups and interactions | 3-4 |
|  |  | (c) Explain how missing data were addressed | 3-4 |
|  |  | (d) If applicable, describe analytical methods taking account of sampling strategy | NA |
|  |  | (e) Describe any sensitivity analyses | NA |
| Results |  |  |  |
| Participants | 13* | (a) Report numbers of individuals at each stage of study—eg numbers potentially eligible, examined for eligibility, confirmed eligible, included in the study, completing follow-up, and analysed | 4 |
|  |  | (b) Give reasons for non-participation at each stage | NA |
|  |  | (c) Consider use of a flow diagram | NA |
| Descriptive data | 14* | (a) Give characteristics of study participants (eg demographic, clinical, social) and information on exposures and potential confounders | 4-6 |
|  |  | (b) Indicate number of participants with missing data for each variable of interest | 4-10 |
| Outcome data | 15* | Report numbers of outcome events or summary measures | 4-10 |
| Main results | 16 | (a) Give unadjusted estimates and, if applicable, confounder-adjusted estimates and their precision (eg, 95% confidence interval). Make clear which confounders were adjusted for and why they were included | 6-10 |
|  |  | (b) Report category boundaries when continuous variables were categorized | NA |
|  |  | (c) If relevant, consider translating estimates of relative risk into absolute risk for a meaningful time period | NA |
| Other analyses | 17 | Report other analyses done—eg analyses of subgroups and interactions, and sensitivity analyses | 11-13 |
| **Discussion** |  |  |  |
| Key results | 18 | Summarise key results with reference to study objectives | 13 |
| Limitations | 19 | Discuss limitations of the study, taking into account sources of potential bias or imprecision. Discuss both direction and magnitude of any potential bias | 13 |
| Interpretation | 20 | Give a cautious overall interpretation of results considering objectives, limitations, multiplicity of analyses, results from similar studies, and other relevant evidence | 13-14 |
| Generalisability | 21 | Discuss the generalisability (external validity) of the study results | 13-14 |
| **Other information** |  |  |  |
| Funding | 22 | Give the source of funding and the role of the funders for the present study and, if applicable, for the original study on which the present article is based | 14 |

\*Give information separately for exposed and unexposed groups.
**Note:** An Explanation and Elaboration article discusses each checklist item and gives methodological background and published examples of transparent reporting. The STROBE checklist is best used in conjunction with this article (freely available on the Web sites of PLoS Medicine at http://www.plosmedicine.org/, Annals of Internal Medicine at http://www.annals.org/, and Epidemiology at http://www.epidem.com/). Information on the STROBE Initiative is available at www.strobe-statement.org.

## REFERENCES

1 Australian Government Department of Health. Coronavirus (COVID-19) at a glance - 8 May 2020. [date accessed: 24th July 2020]. Available from: https://www.health.gov.au/resources/publications/coronavirus-covid-19-at-a-glance-8-may-2020.

2 World Health Organisation. WHO Coronavirus Disease (COVID-19) Dashboard. [date accessed: 24th July 2020]. Available from: https://covid19.who.int/.

3 Reuters in Kuala Lumpur. Global condom shortage looms as coronavirus shuts down production. The Guardian 2020. Available from: https://www.theguardian.com/world/2020/mar/27/global-condom-shortage-coronavirus-shuts-down-production

4 Marie Stopes Australia. Situational Report: Sexual and Reproductive Health Rights in Australia, Updated 8 May 2020. 08 May 2020 [date accessed: 22 May 2020]. Available from: https://resources.mariestopes.org.au/SRHRinAustralia.pdf.

5 Plan International. Periods in a pandemic: menstrual hygine management in the time of COVID-19. 2020 [date accessed: 19 June 2020]. Available from: https://www.plan.org.au/-/media/plan/documents/learn/publications/periods-in-a-pandemic.pdf.

6 Winter, C. Demographer warns of coronavirus ‘ripple effects’, predicts drop in birthrate and taxpayers. Australian Broadcasting Company (ABC) News (online) 2020. Available from: https://www.abc.net.au/news/2020-04-27/demographer-warns-of-the-missing-children-of-covid-19/12187262

7 QSR International Pty Ltd, NVivo qualitative data analysis software. 2012, QSR International Pty Ltd.

8 Braun, V., Clarke, V., and Terry, G., Thematic Analysis, in Qualitative Research in Clinical and Health Psychology, P. Rohleder and A.C. Lyons, Editors. 2015, Palgrave Macmillan.

9 Sexual Health Unit The University of Melbourne. Sexual and Reproductive Health during COVID-19 (coronavirus). [date accessed: 5th August 2020]. Available from: https://mspgh.unimelb.edu.au/research-groups/centre-for-epidemiology-and-biostatistics-research/sexual-health/sexual-and-reproductive-health-during-covid-19-coronavirus.

10 Wilkins, R., Who’s hit hardest by the economic effects of COVID-19?, Melbourne Institute: The University of Melbourne. Available from: https://melbourneinstitute.unimelb.edu.au/_data/assets/pdf_file/0006/3387039/ri2020n10.pdf

11 Family Planning New South Wales, Contraception in Australia: 2005-2018, FPNSW: Ashfield, Sydney. Available from: https://www.fpnsw.org.au/sites/default/files/assets/Contraception-in-Australia_2005-2018_v20200716.pdf

12 Coombe, J., Harris, M.L., and Loxton, D. Examining long-acting reversible contraception non-use among Australian women in their 20’s: Findings from a qualitative study. Culture, Health & Sexuality, 2019. 21;7:822–36.

13 The Royal Australian and New Zealand College of Obstetricians and Gynaecologists. A message for pregnant women and their families. 2020 [date accessed: 30th July 2020]. Available from: https://ranzcog.edu.au/statements-guidelines/covid-19-statement/information-for-pregnant-women.

14 Loxton, D., Forder, P., Townsend, N., et al., ALSWH COVID-19 Survey Report 4: Survey 4, 10 June 2020, Available from: https://www.alswh.org.au/images/content/reports/ALSWH_COVID-19_Survey_4_Report_Final.pdf

15 Gender Equity Victoria. The balance between safety and support essential for the mental health of expectant mothers. [date accessed: 30th July 2020]. Available from: https://www.genvic.org.au/media-releases/the-balance-between-safety-and-support-essential-for-the-mental-health-of-expectant-mothers/.

16 Australian Broadcasting Company (ABC) News. Scott Morrison says panic buying driven by coronavirus lockdown fears is ‘ridiculous’ and ‘un-Australian’. Wednesday 18th March 2020 [date accessed: 17 July 2020]. Available from: https://www.abc.net.au/news/2020-03-18/coronavirus-panic-buying-pm-tells-people-to-stop-hoarding/12066082.

17 Minister for Health: Greg Hunt. Media Statement: Continuous care with telehealth stage seven. [date accessed: 17th July 2020]. Available from: https://www.greghunt.com.au/continuous-care-with-telehealth-stage-seven/.

18 Marie Stopes Australia. Medicare rollback will limit sexual & reproductive health access. [date accessed: 17 July 2020]. Available from: https://www.mariestopes.org.au/your-choices/medicare-rollback-limit-access/.

19 Australian Government Department of Education Skills and Employment. Early Childhood Education and Care COVID-19 Frequently Asked Questions. [date accessed: 30th July 2020]. Available from: https://www.dese.gov.au/covid-19/childcare/childcare-faq#:~:text=The%20Early%20Childhood%20Education%20and%20Care%2ORelief%20Package%20(Relief%20Package,end%20on%2028%20June%202020.

20 Sheridan, A. The case for maintaining free childcare - Australia’s “Pink Collar Recession”. The Canberra Times 2020. Available from: https://www.canberratimes.com.au/story/6823171/the-case-for-maintaining-free-childcare-australias-pink-collar-recession/

